# Wastewater surveillance using ddPCR reveals highly accurate tracking of Omicron variant due to altered N1 probe binding efficiency

**DOI:** 10.1101/2022.02.18.22271188

**Authors:** Melissa K. Schussman, Adelaide Roguet, Angela Schmoldt, Brooke Dinan, Sandra L. McLellan

## Abstract

Wastewater surveillance for SARS-CoV-2 is being used worldwide to understand COVID-19 infection trends in a community. We found the emergence and rapid timeline for dominance of the Omicron variant was accurately reflected in wastewater when measured with droplet digital (dd)PCR. We were able to distinguish Omicron from the circulating Delta variant because Omicron has a mutation in the N1 probe binding region that diminished the fluorescent signal within individual droplets. The ddPCR platform may be advantageous for wastewater surveillance since analysis of the data can segregate fluorescent signals from different individual templates. In contrast, platforms such as qPCR that rely solely on the intensity of fluorescence for quantification would not distinguish a subset of variants with mutations affecting the reaction and could underestimate SARS-CoV-2 concentrations. The proportion of Omicron in wastewater was tightly correlated to clinical cases in five cities and provided a higher resolution timeline of appearance and dominance (>75%) than sequenced clinical samples, which were limited in less populated areas. Taken together, this work demonstrates wastewater is a reliable metric for tracking SARS-CoV-2 at a population level.

## Main text

Wastewater surveillance has gained traction as a public health tool to track COVID-19 infections in the community. In just two years, methods have been developed to concentrate and quantify the SARS-CoV-2 virus from wastewater (1, 2). The rapid emergence of variants of concern forces the wastewater monitoring scientific community to continuously adapt their methods. The Omicron variant is unique in that it contains a mutation in the N1 gene that corresponds to the N1 probe binding site of the CDC assay (3). Digital (d)PCR and droplet digital (dd)PCR platforms provide data for individual molecules and can reveal a decreased but positive fluorescence intensity in discrete reactions. Here we show how wastewater surveillance using ddPCR was able to accurately track the Omicron variant emergence and fixation in five communities in Wisconsin.

## The study

We quantified SARS-CoV-2 concentrations in influent samples from seven wastewater treatment plants (WWTPs) in five communities as part of our ongoing Wisconsin SARS-CoV-2 wastewater surveillance program (4). This program has been part of the National Wastewater Surveillance System since August 2020 (5). Concentration and quantification methods for our lab have been described previously (6) and are detailed in the supplemental text S1. Detection of the Omicron variant was derived from ddPCR droplet data, which was processed using the QuantaSoft Software, version 1.7, for the Bio-Rad QX200 Droplet Digital System (Bio-Rad, Hercules, CA). To quantify the concentration of the Omicron variant in each sample, the lasso threshold adjustment tool was used to reclassify the cluster delineation in the 2D amplitude scatterplots. To ensure the correct amplitude was being associated with each respective variant, we included an Omicron and Delta residual clinical sample diluted 1:100 in addition to two 1:8 diluted Exact Diagnostics SARS-CoV-2 standards (Bio-Rad).

Over the course of late November through December 2021, we found that SARS-CoV-2 levels steadily increased and the N1 signal in ddPCR could be separated into two distinct clusters, one with lower fluorescence than expected based on the N1 standard (**Figure 1**). The Omicron variant has a C to U mutation at position 28,311 in the SARS-CoV-2 genome, which corresponds to the 3rd nucleotide position on the 5′ end of the probe for the CDC N1 PCR assay (7). The loss of fluorescent signal is likely due to inefficient exonuclease activity of the Taq polymerase on the 5′ end of the probe which releases the fluorophore from the quencher. The effects of probe mismatches on florescent signal has been previously reported (8). The probe is impacted more by mutations nearer the 5′ end, in contrast to primers, where mismatches in the 3′ end are more likely to affect the extension activity of the polymerase (9). Overall, mutations in the probe rather than the primers are more likely to result in reduced quantification (10). One preprint reported the Omicron mutation in the N1 probe of the CDC assay did not perturb N1 detection, but did note a slightly reduced cycle threshold (Ct) value for the N1 target (11). This study was focused on diagnosis of positive clinical samples and not reporting quantitative values in wastewater where accurate quantification is necessary.

**Figure 1.**
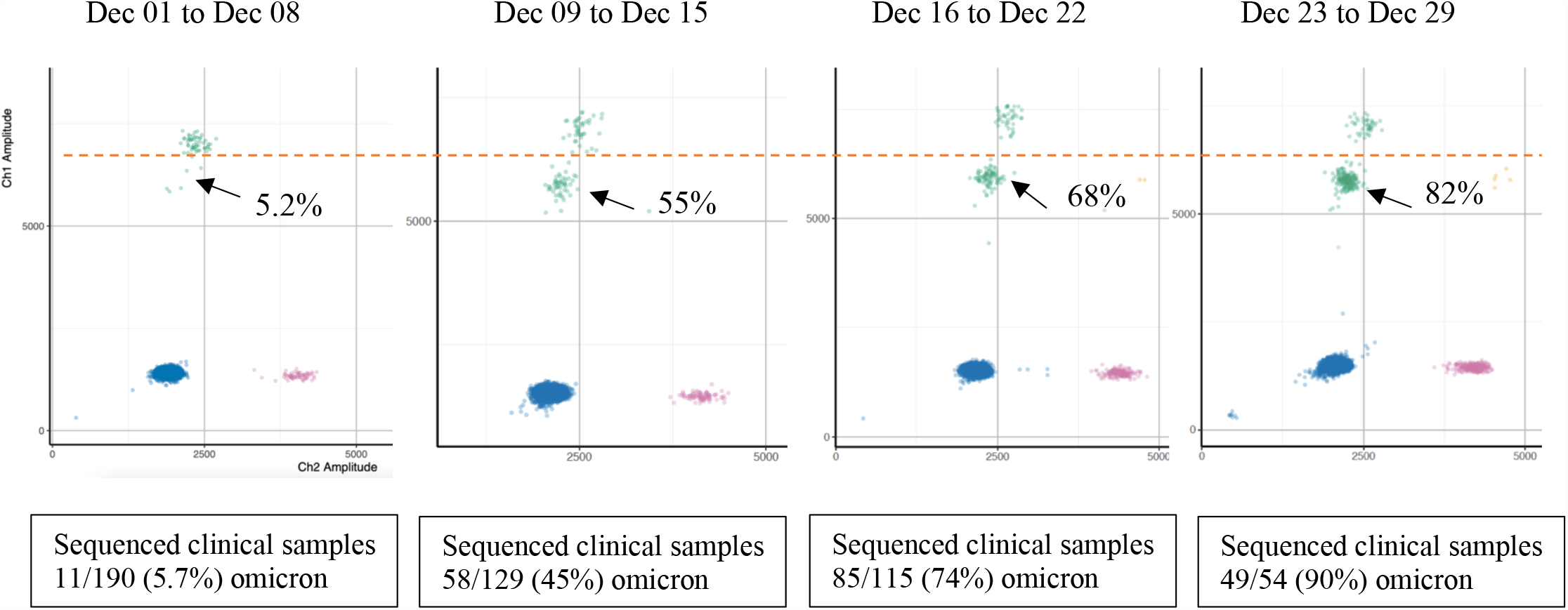
Wastewater N1N2 multiplex ddPCR results in the Jones Island WWTP, Milwaukee, Wisconsin throughout December 2021 during the Omicron variant surge. A reduced fluorescence in the N1 signal in Channel 1 (green droplets) resulted in two clouds and was indicative of Omicron. N2 signal is in Channel 2 (pink droplets). Channel 1 and 2 fluorescence in the same droplet indicate both N1 and N2 target are present (yellow droplets) Negative droplets are blue. Note the steady increase (∼10 fold) in NI and N2 signal over 4 weeks. Clinical patient samples sequenced from Milwaukee County each week are indicated below graph (data from GISAID).

We validated the accuracy of quantifying the Omicron variant using the N1 cloud splits with the TaqMan SARS-CoV-2 Mutation Panels S.P681R.CCT.CGT (Delta) and S.P681H.CCT.CAT (Omicron) (ThermoFisher Scientific, Waltham, MA) in two WWTPs. A significant positive correlation was observed between the specific mutation assay and N1 cloud split quantification for Omicron (Spearman’s rank correlation, rho = 0.845, p < 0.0005) and Delta variants (Spearman’s rank correlation, rho = 0.785, p < 0.0005). The trends in the variant concentrations mirrored each other (Supplemental Figure 1), but overall the N1 assay was more efficient (i.e. higher number of droplets) than the specific mutation assays.

We also compared the proportion of Omicron and Delta in wastewater with sequencing results from clinical samples available in GISAID (12) for five cities serviced by seven WWTPs (**Figure 2**). We assumed that county-level data would be a proxy for different cities within the county. There was good agreement between WWTPs in the same city or county, and in all cases, the WWTP serving the larger population detected omicron earlier. In 6 of 7 WWTPs, Omicron was detected >limit of detection (LOD) prior to, or on the same date as, the first clinically confirmed case of Omicron (**Table 1**). Our first sample indicating a second N1 cluster above the limit of detection (3 positive droplets) was a sample from November 21, 2021, one day prior to the first diagnosed clinical Omicron sample in the US.

**Figure 2.**
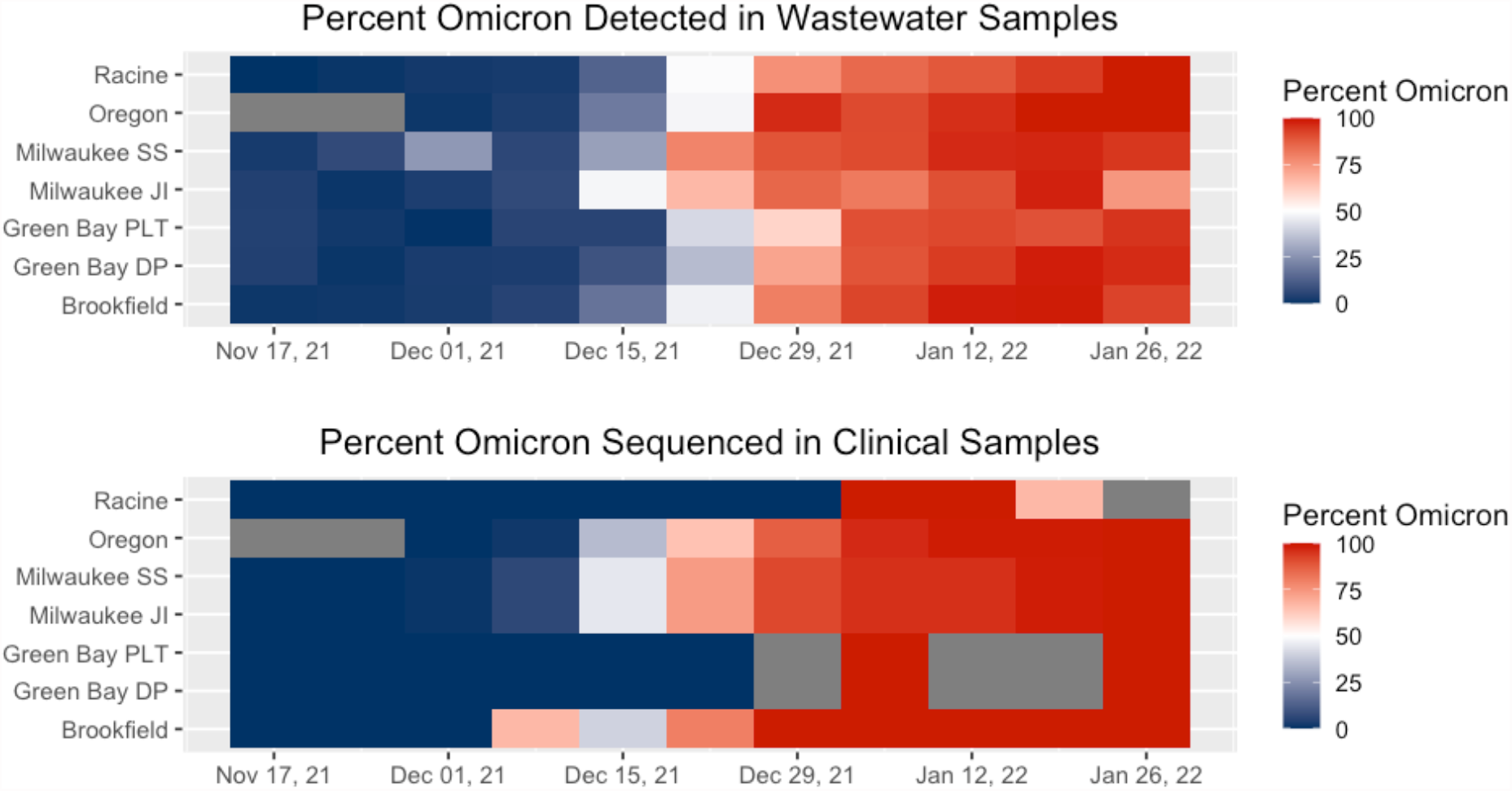
Heat map of percentage of Omicron in (a) WWTP samples determined by ddPCR and (b) clinical samples determined by sequencing as reported in GISAID. In general, the WWTP samples showed a more gradual progression. Gray bars indicate no clinical samples were sequenced in the counties in which those cities are located.

**Table 1.** Dates of first detection of Omicron by whole genome sequencing of clinical samples and routine analysis of ddPCR of wastewater samples.

| WWTP | County | First clinical | First >LOD wastewater | Days from first >LOD to 75% Omicron | Population |
| --- | --- | --- | --- | --- | --- |
| Racine | Racine | 5-Jan-22 | 5-Dec-21 | 28 | 139000 |
| Oregon | Dane | 12-Dec-21 | 8-Dec-21 | 29 | 10000 |
| Milwaukee SS | Milwaukee | 1-Dec-21 | 21-Nov-21 | 35 | 615934 |
| Milwaukee JI | Milwaukee | 1-Dec-21 | 5-Dec-21* | 21 | 470007 |
| Green Bay PLT | Brown | 30-Dec-21 | 28-Nov-21 | 39 | 189000 |
| Green Bay DP | Brown | 30-Dec-21 | 6-Dec-21 | 27 | 43000 |
| Brookfield | Waukesha | 7-Dec-21 | 28-Nov-21 | 30 | 51000 |

\*Milwaukee JI 17-Nov-21 sample was run in three ddPCR wells to increase sensitivity and found to be >LOD. All data in Table 1 represents measurements from a single well of ddPCR, which is used for routine monitoring.

This date is identical to east and west coast wastewater detection (13), but is noteworthy as variants of concern are not usually first observed in Midwest states. Retrospective analysis using multiple ddPCR reactions to increase sensitivity for the 17-Nov-21 Milwaukee JI sample demonstrated Omicron was present in wastewater one week prior.

When comparing our wastewater data to sequenced clinical samples, we saw a later onset of Omicron in clinical samples, with a more rapid spike in percentage. In addition, there were various instances where no clinical samples were sequenced for that county. Further, the low number of clinical samples sequenced for less populated parts of the state might not accurately portray proportions. For example, 23 clinical samples were sequenced in Brown County and 37 in Waukesha for the entire month of December. Because clinical sequencing may not be equally resourced across a state, the results of wastewater testing are expected to be more consistent. Further, sequencing of clinical samples might be biased toward testing for variants in samples from vaccinated individuals or in samples with S gene target failure, which can be an indicator of specific variants (14).

## Conclusions

Here we show that wastewater accurately captured the emergence of the Omicron variant, which coincided with a steep increase in SARS-CoV-2 wastewater concentrations. Since methods for SARS-CoV-2 are relatively new, it has been critical to have a diverse array of approaches. Recent evaluations show the ddPCR platform has higher sensitivity and less susceptibility to inhibition the qPCR (15). In this work, we also show the distinct advantage of segregating the intensity of fluorescent signals from individual templates. Platforms such as qPCR that rely solely on the intensity of fluorescence of the probe for quantification could underestimate SARS-CoV-2 concentrations when there is a subset of variants with mutations affecting the reaction. It is unclear how other methods of quantification are affected by the mutation in the N1 probe, but this warrants further investigation. As new variants emerge, ddPCR may be a preferred platform for wastewater surveillance since reduced PCR efficiency associated with a critical mutation in the probe (or primers) can be flagged in the data, whereas with other platforms, these anomalies may be harder to recognize.

This work adds to the growing body of evidence that wastewater is a cost-effective and accurate population-level measure that provides the same level or more information as traditional public health metrics. Two of the cities we analyzed had a very low number of sequenced clinical samples, whereas the timing of the Omicron appearance and dominance was easily detected in wastewater. This proof of concept adds confidence to public health officials that are considering using wastewater data (16). Public health entities are beginning to use SARS-CoV-2 levels in wastewater for allocating testing resources and evaluating possible irregularities in traditional surveillance (5), and as at-home testing becomes more commonplace or clinical sample sequencing declines, wastewater data may become a primary metric they rely upon.

## Data Availability

All data produced in the present study are available upon reasonable request to the authors

## Funding

This work was funded by a grant awarded to the Wisconsin State Laboratory of Hygiene (subcontract to UW-Milwaukee) from the Wisconsin Department of Health Services (DHS) administered, CDC-funded, ELC (Epidemiology and Laboratory Capacity) Enhanced Detection Expansion program.

## Acknowledgments

We thank Jonathan Meiman and Nathan Kloczko from Wisconsin DHS and the wastewater team at the Wisconsin State Laboratory of Hygiene for insightful discussion and knowledge sharing.

## Notes

### Competing Interest Statement

The authors have declared no competing interest.

