## Supplemental Material for "Wastewater surveillance using ddPCR reveals highly accurate tracking of Omicron variant due to altered N1 probe binding efficiency"

#### Methodology

SARS-CoV-2 concentration and variant analysis includes a total of 165 samples that were collected between November 14, 2021, and January 31, 2022. Flow-weighted composite samples were collected twice per week over a 24-hour period from seven WWTP's: Brookfield, Green Bay DP, Green Bay PLT, Milwaukee JI, Milwaukee SS, Oregon, and Racine. Samples were filtered and according to Feng et al. 2021. Extraction was performed using a custom Maxwell® HT Environmental TNA Kit (Promega, Madison, WI, USA) on an automated instrument KingFisher Flex (Thermo Fisher Scientific, Waltham, MA, USA).

The ddPCR assay probes used to quantify N1 and N2 regions of SARS-CoV-2, as well as Delta and Omicron variant are shown in Table S1. All ddPCR assays were performed in 22-µL reaction mixtures using the one-step RT-ddPCR Advanced Kit for Probes (Bio-Rad, Hercules, CA, USA), amplified using the Mastercycler pro (Eppendorf, Hamburg, Germany), and read using the Bio-Rad QX200 Droplet Digital System as according to Feng et al. 2021.

**Table S1** List of the ddPCR assays used

| Target | Primer/Probe Sequence (5'-3') | Supplier | Ref |
|---|---|---|---|
| SARS-CoV-2 N1 | F: GACCCCAAAATCAGCGAAAT<br>R: TCTGGTTACTGCCAGTTGAATCTG<br>P: FAM/ACCCCGCAT/ZEN/TACGTTTGGTGGACC/lABkFQ | Eurofins<br>Eurofins<br>IDT | [1] |
| SARS-CoV-2 N2 | F: TTACAAACATTGGCCGCAAA<br>R: GCGCGACATTCCGAAGAA<br>P: HEX/ACAATTTGCCCCCAGCGCTTCAG/BHQI and HEX/ACAATTTGC/ZEN/CCCCAGCGCTTCAG/lABkFQ | Eurofins<br>Eurofins<br>IDT | [1] |
| Delta (P681R) | Reporter 1 dye: VIC, Reporter 1 Quencher: NFQ,<br>Reporter 2 dye: FAM, Reporter 2 quencher: NFQ<br>CTCAGACTAATTCTC[C/A]TCGGCGGGCACGTAG | Applied Biosystems | [3] |
| Omicron (P681H) | Reporter 1 dye: VIC, Reporter 1 Quencher: NFQ,<br>Reporter 2 dye: FAM, Reporter 2 quencher: NFQ,<br>CTCAGACTAATTCTC[C/G]TCGGCGGGCACGTAG | Applied Biosystems | [3] |

*ddPCR primers and probes at a final concentration of 900 nM and 250 nM, respectively.

*Reporter 2 dye targets Mutant of TaqMan SARS-CoV-2 Mutation Panels.

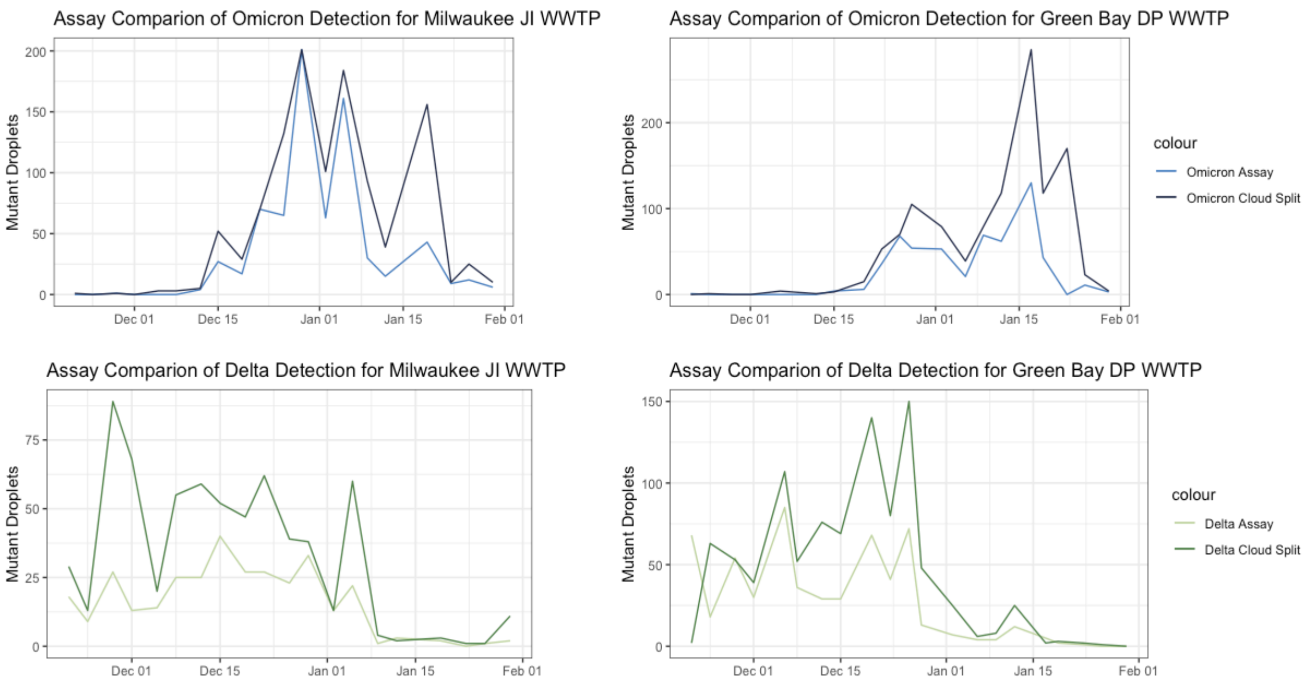


**Figure S1** Comparison of the quantification of Omicron and Delta mutant droplets using the TaqMan SARS-CoV-2 Mutation panels and N1 probe cloud split methods. Shapiro-Wilk normality test and Spearman's rank correlation analysis performed in R-Studio version 1.4.0013.

### References

[1] CDC, "2019-Novel Coronavirus ( 2019-nCoV ) Real-time rRT-PCR Panel Primers and Probes Note," *Div. Viral Dis. Centers Dis. Control Prev.*, 2020.

[2] Feng, Shuchen, Adelaide Roguet, Jill S. McClary-Gutierrez, Ryan J. Newton, Nathan Kloczko, Jonathan G. Meiman, and Sandra L. McLellan. 2021. "Evaluation of Sampling, Analysis, and Normalization Methods for SARS-CoV-2 Concentrations in Wastewater to Assess COVID-19 Burdens in Wisconsin Communities." *ACS ES&T Water* 1 (8): 1955–65.

[3] "TaqMan SARS-CoV-2 Mutation Panel." Applied Biosystems by Thermo Fisher Scientific, https://www.thermofisher.com/us/en/home/clinical/clinical-genomics/pathogen-detection-solutions/real-time-pcr-research-solutions-sars-cov-2/mutation-panel.html#menu5
